# Intravenous thrombolysis prior to endovascular treatment in posterior circulation occlusions; a MR CLEAN Registry study

**DOI:** 10.1101/2023.05.16.23290075

**Authors:** R.R.M.M. Knapen, F.A.V. Pirson, L.C.M. Langezaal, J. Brouwer, C.B. Majoie, B.J. Emmer, J.A. Vos, P.J. van Doormaal, A.J. Yoo, A.A.E. Bruggeman, G.J. Lycklama à Nijeholt, C. van der Leij, R.J. van Oostenbrugge, W.H. van Zwam, W.J. Schonewille, MR CLEAN Registry Investigators.

## Abstract

**Background and aims:** The effectiveness of intravenous thrombolysis (IVT) prior to endovascular treatment (EVT) has been investigated in randomized trials and meta-analyses. These studies mainly concerned anterior circulation occlusions. We aimed to investigate clinical, technical, and safety outcomes of IVT prior to EVT in posterior circulation occlusions in a nationwide registry.

**Methods:** Patients were included from the MR CLEAN Registry: a nationwide, prospective, multicenter registry of patients with acute ischemic stroke (AIS) due to a large intracranial vessel occlusion receiving EVT between 2014 and 2019. All patients with a posterior circulation occlusion were included. Primary outcome was a shift towards better functional outcome on the modified Rankin scale (mRS) at 90 days. Secondary outcomes were favorable functional outcome (mRS 0-3), occurrence of symptomatic intracranial hemorrhages (sICH), successful reperfusion (eTICI≥2B), first-attempt successful reperfusion, and mortality at 90 days. Regression analyses with adjustments based on univariate analyses and literature were applied.

**Results:** A total of 248 patients were included, who received either IVT (n=125) or no IVT (n=123) prior to EVT. Results show no differences in a shift on the mRS (acOR:1.04, 95%CI:0.61-1.76). Although sICH occurred more often in the IVT group (4.8% versus 2.4%), regression analysis did not show a significant difference (aOR:1.65, 95%CI:0.33-8.35). Successful reperfusion, favorable functional outcome, first-attempt successful reperfusion, and mortality did not differ between patients treated with and without IVT.

**Conclusions:** We found no significant differences in clinical, technical and safety outcomes between patients with a large vessel occlusion in the posterior circulation treated with or without IVT prior to endovascular therapy. Our results are in line with the literature on the anterior circulation.

## Introduction

Intravenous thrombolysis (IVT) prior to endovascular treatment (EVT) is recommended in all patients with ischemic stroke due to an intracranial large vessel occlusion in the anterior circulation within 4.5 hours after symptom onset.(1) Although treatment with IVT between 4.5 and 9 hours may be considered in the presence of a mismatch on CT perfusion in the anterior circulation, there is no consensus about the indication for IVT prior to EVT in this late time window.(1)

Recent meta-analyses and randomized clinical trials (RCTs) found no superiority or non-inferiority in functional outcome and mortality at 90 days between patients with a large vessel occlusion (LVO) treated with and without IVT prior to EVT.(2–8) These studies mainly concerned patients with anterior circulation occlusions.

The BEST, BASICS, ATTENTION, and BAOCHE trials are RCTs on the effectiveness of EVT in patients with a basilar artery occlusion. (9–12) ATTENTION and BAOCHE showed a beneficial effect of EVT in patients treated within 12 hours and between 6-24 hours of symptom onset, respectively. However, no RCTs are available on the effectiveness of IVT in posterior circulation occlusions.(13) Two meta-analyses, based on cohort studies, showed lower incidences of intracranial hemorrhage in patients treated with IVT alone for posterior circulation stroke as compared to anterior circulation stroke.(14, 15) In patients with posterior circulation stroke compared to anterior circulation stroke treated with IVT, but without EVT, higher mortality rates were found.(15) When patients were treated with IVT prior to EVT, sICH rates were comparable and mortality rates were higher in the posterior circulation occlusion as compared to anterior circulation occlusion.(15)

Since the available data from the literature is limited, our study aimed to investigate the outcomes of patients with posterior circulation occlusion treated with EVT, with or without prior IVT in a large nationwide registry (MR CLEAN Registry).(16)

## Methods

### Design and participants

Patients were included from the MR CLEAN (Multicenter Randomized Clinical Trial of Endovascular Treatment for Acute Ischemic Stroke in the Netherlands) Registry: a prospective, observational study in all EVT performing centers (n=18) in the Netherlands. The registry included patients treated with EVT for acute ischemic stroke due to large vessel occlusion between March 2014 and December 2018. The MR CLEAN Registry study protocol was evaluated by the medical ethics committee of the Erasmus University Medical Center and permission was granted to carry out the study as a registry. The need for obtaining informed consent was waived. For the current study, the following inclusion criteria were used: age ≥ 18 years, NIHSS ≥ 2; occlusion in the posterior circulation confirmed by CT-angiography (CTA). Patients in whom no intracranial access was obtained were excluded.

### Outcome measures

The primary outcome was the modified Rankin Scale score at 90 days follow-up, ranging from 0 (no disability) to 6 (death). Secondary outcomes were favorable functional outcome (defined as mRS 0-3), functional independent outcome (defined as mRS 0-2), and the National Institute of Health Stroke Scale (NIHSS) score at 24-48 hours. Technical outcomes included procedure duration (defined as groin puncture to reperfusion), first-attempt successful reperfusion and successful reperfusion. Safety outcomes were the occurrence of symptomatic intracranial hemorrhages (sICH) within 3 days after EVT, mortality at 90 days, and serious adverse events (e.g. stroke progression and pneumonia).

### Imaging assessment

Intracranial hemorrhage was defined as symptomatic when the patient had neurological deterioration (at least 4 points increase on the NIHSS) in combination with a hemorrhage (according to the Heidelberg criteria), which was related to the clinical deterioration. An adverse event committee evaluated the medical reports and imaging to determine a sICH.

Recanalization status was scored on digital subtraction angiography (DSA) according to the extended Thrombolysis in Cerebral Ischemia (eTICI) by an independent core laboratory. This core laboratory consisted of eight interventional radiologists or neuroradiologists, all blinded to the clinical findings. The eTICI ranges from 0 (no reperfusion) to 3 (complete reperfusion). In this study, successful reperfusion was defined as eTICI≥2B (50-90% reperfusion of affected area), excellent reperfusion as eTICI 3, and first-attempt successful recanalization as eTICI≥2C (90-99% reperfusion of affected area) in combination with one attempt. When only a DSA was performed because of recanalization, it was registered as early recanalization. When the DSA was made in only one direction, the maximum eTICI score was set at 2A. The posterior circulation Acute Stroke Prognosis Early Computed Tomography Score (PC-ASPECTS) was scored on non-contrast CT (NCCT), while the posterior circulation collateral score (PC-CS) was scored on baseline CTA by the core laboratory.

### Statistical analysis

Baseline characteristics were presented using descriptive statistics. Dichotomous and ordinal parameters were compared using Pearson’s chi-squared test or Fisher’s exact test. Continuous variables were tested using independent-samples t-test or Mann-Whitney U test, after checking for the normality using histograms.

For the primary outcome, a multivariable ordinal logistics regression model was used to compare the use of IVT for a one-step shift on the mRS score at 90 days follow-up. Continuous variables were checked on normality of distribution of the residuals using Q-Q plots. When no normality was seen, the variable was transformed using a natural logarithm. After exponentiating the regression coefficient, relative percentages were calculated using the following formula: (exponentiate(coefficient) -1) *100%. Adjusted (a) odds ratios (OR) or beta estimates with 95% confidence intervals were used to present the regression model results.

All regression models were adjusted for potential confounders: age, sex, baseline NIHSS score, pre-mRS score (dichotomized 0-2 versus 3-5), diabetes mellitus, hypertension in patients’ history, systolic blood pressure when entering the hospital, the use of anticoagulation medication, the collaterals at CTA baseline, and the time between estimated large vessel occlusion and groin puncture. These confounders were chosen based on univariate analyses complemented with parameters observed in previous literature. All analyses were performed using RStudio (version 2022.07.2). The alpha was set at 5%.

### Missing values

Original data were used for the descriptive analyses, whereas multiple imputations were used for the missing data before conducting the regression analyses. The complete list of variables used for imputation is described in Supplemental 1.

## Subgroup analyses

An interaction term was calculated to assess the interaction between occlusion location and IVT on the mRS score at 90 days. Subgroup analyses were added exploratory. The same variables for adjustment were used as for the primary analysis, regardless of the group sizes.

The corresponding author had full access to all the data in this study and takes responsibility for its integrity and the data analysis. Source data will not be made available because of legislative issues on patient privacy. Detailed statistical analyses and analytic methods will be made available on reasonable request to the corresponding author. This study was conducted using the STROBE guidelines.

## Results

### Baseline characteristics

A total of 5768 patients were included in the MR CLEAN Registry, of which 264 patients had a posterior circulation occlusion. After applying the in- and exclusion criteria, a total of 248 patients were analyzed in the current study (Figure 1). Patients with IVT less often used anticoagulation prior to EVT, had lower pre-mRS scores, had faster onset to groin puncture times and more often showed early recanalization compared to the patients treated without IVT (Table 1).

**Figure 1.**
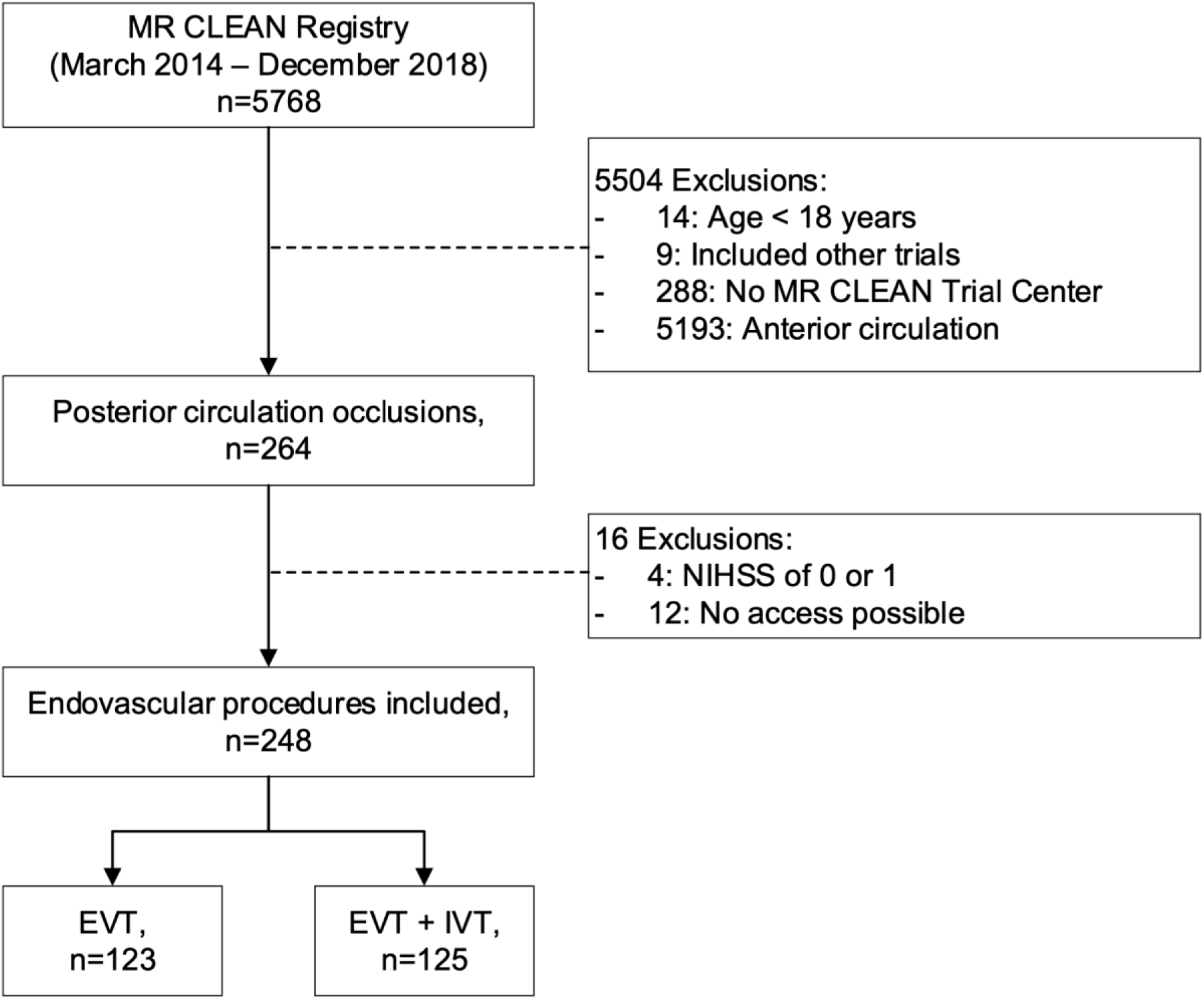
**Flow-chart of included patients in this study.** NIHSS, National Institutes of Health Stroke Scale; EVT, endovascular treatment; IVT, intravenous thrombolysis.

**Figure 2.**
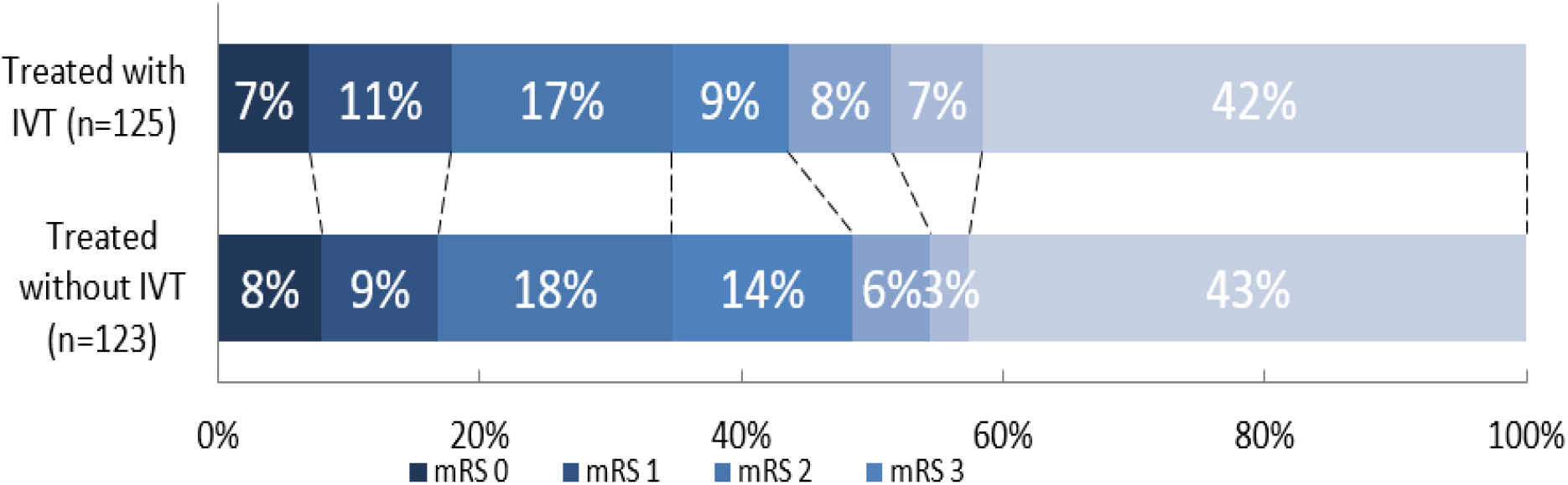
Distribution of the modified Rankin Scale. Multiple logistic regression with adjustment showed no significant difference between patients treated with intravenous thrombolysis prior endovascular treatment compared to patients treated with IVT prior EVT (adjusted common odds ratio 1.04 (95%CI:0.61 – 1.76)). IVT, intravenous thrombolysis; mRS, modified Rankin Scale.

**Table 1.**
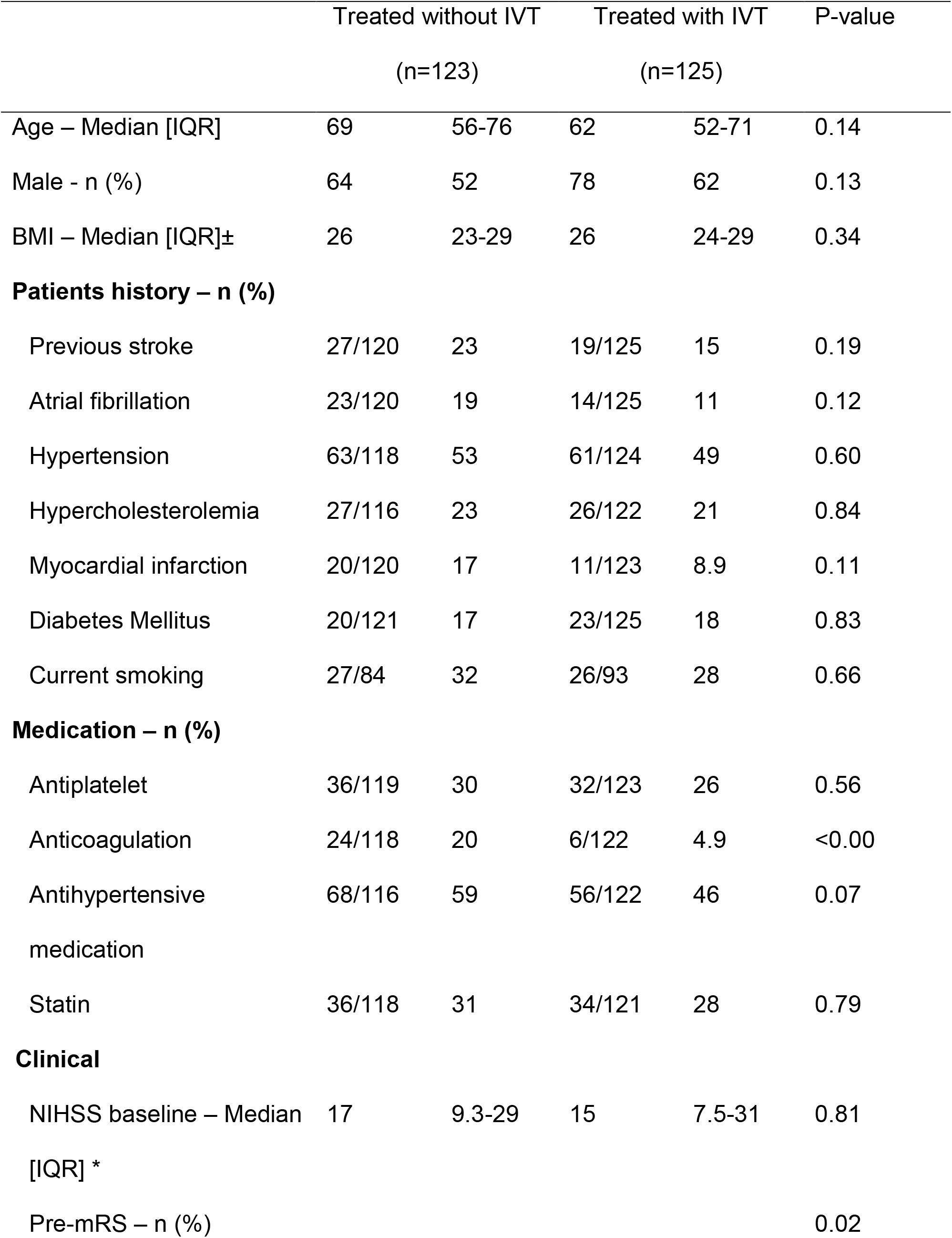
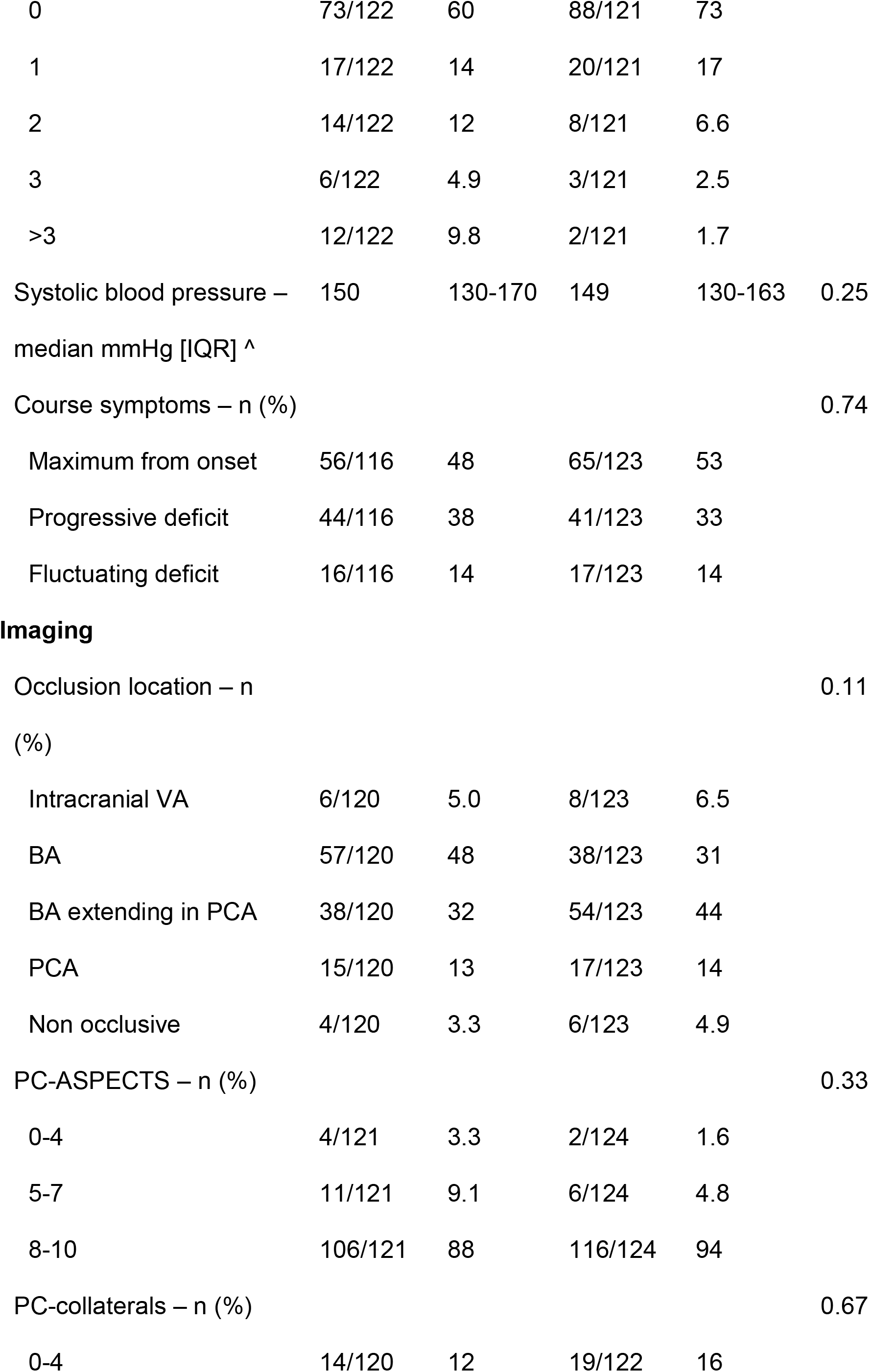
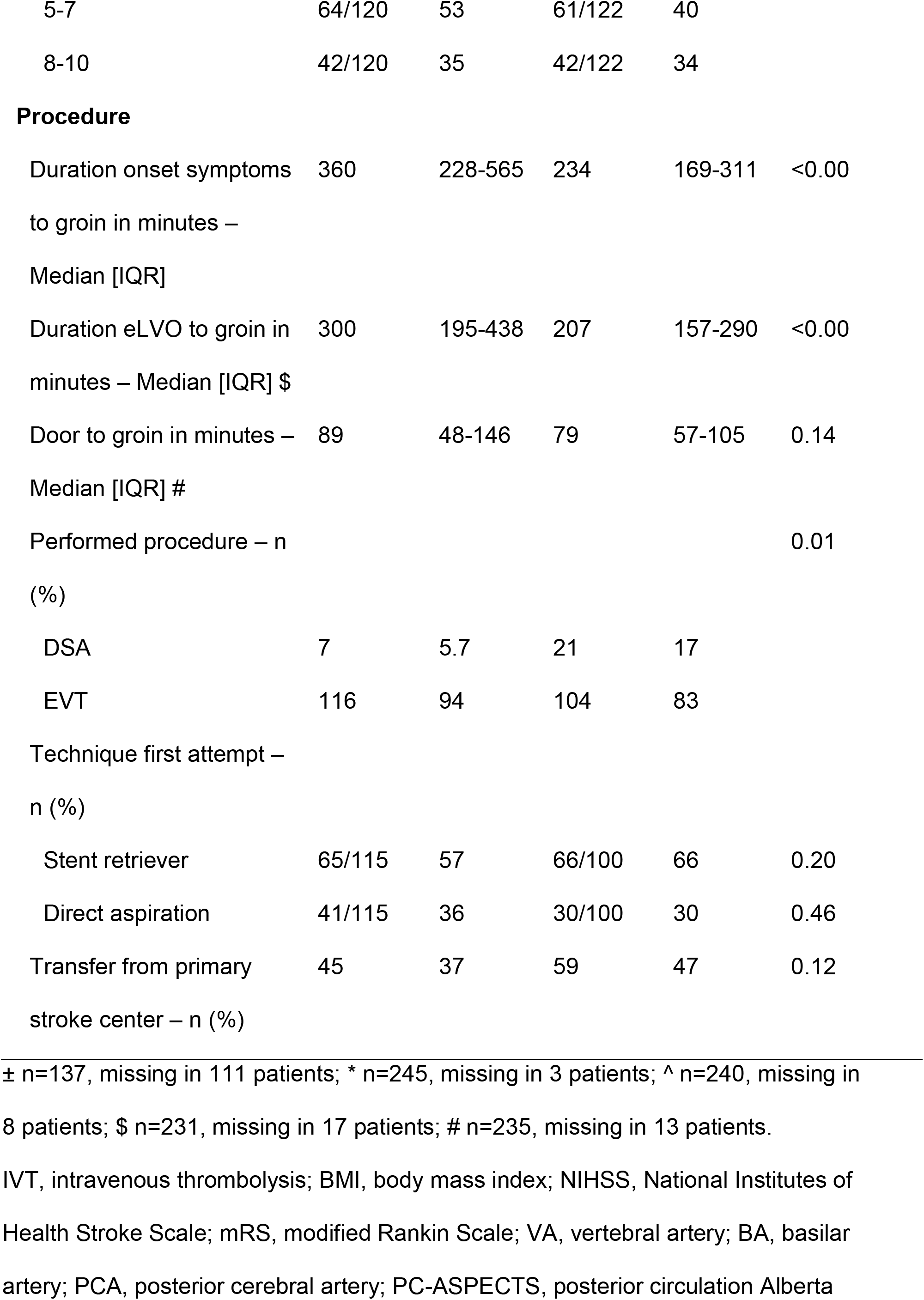
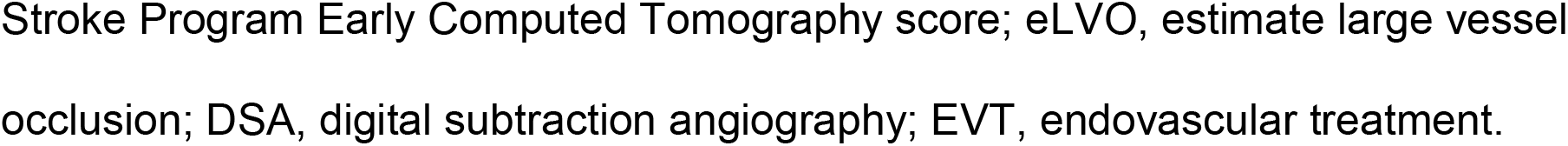

**Table 2.**
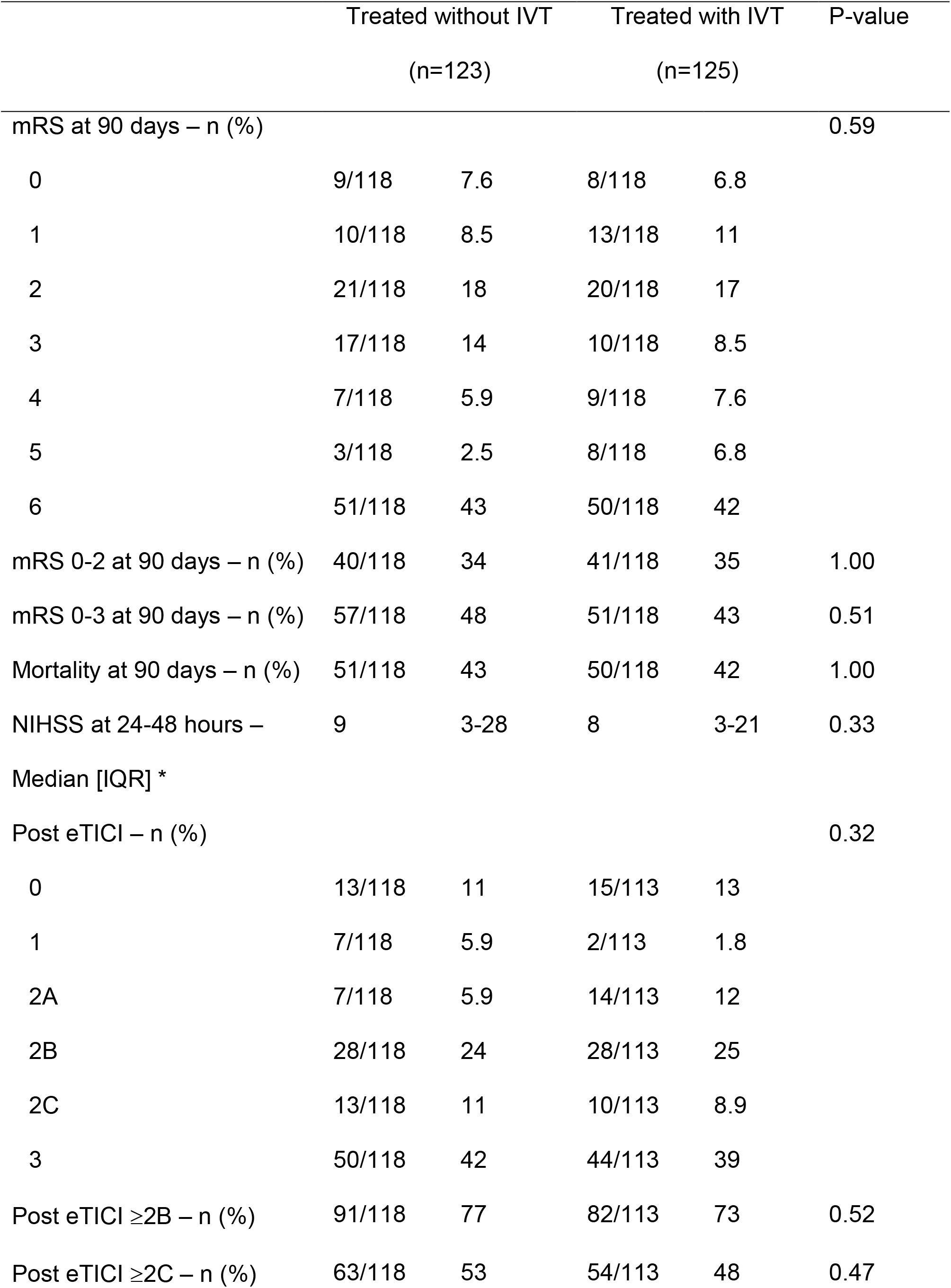
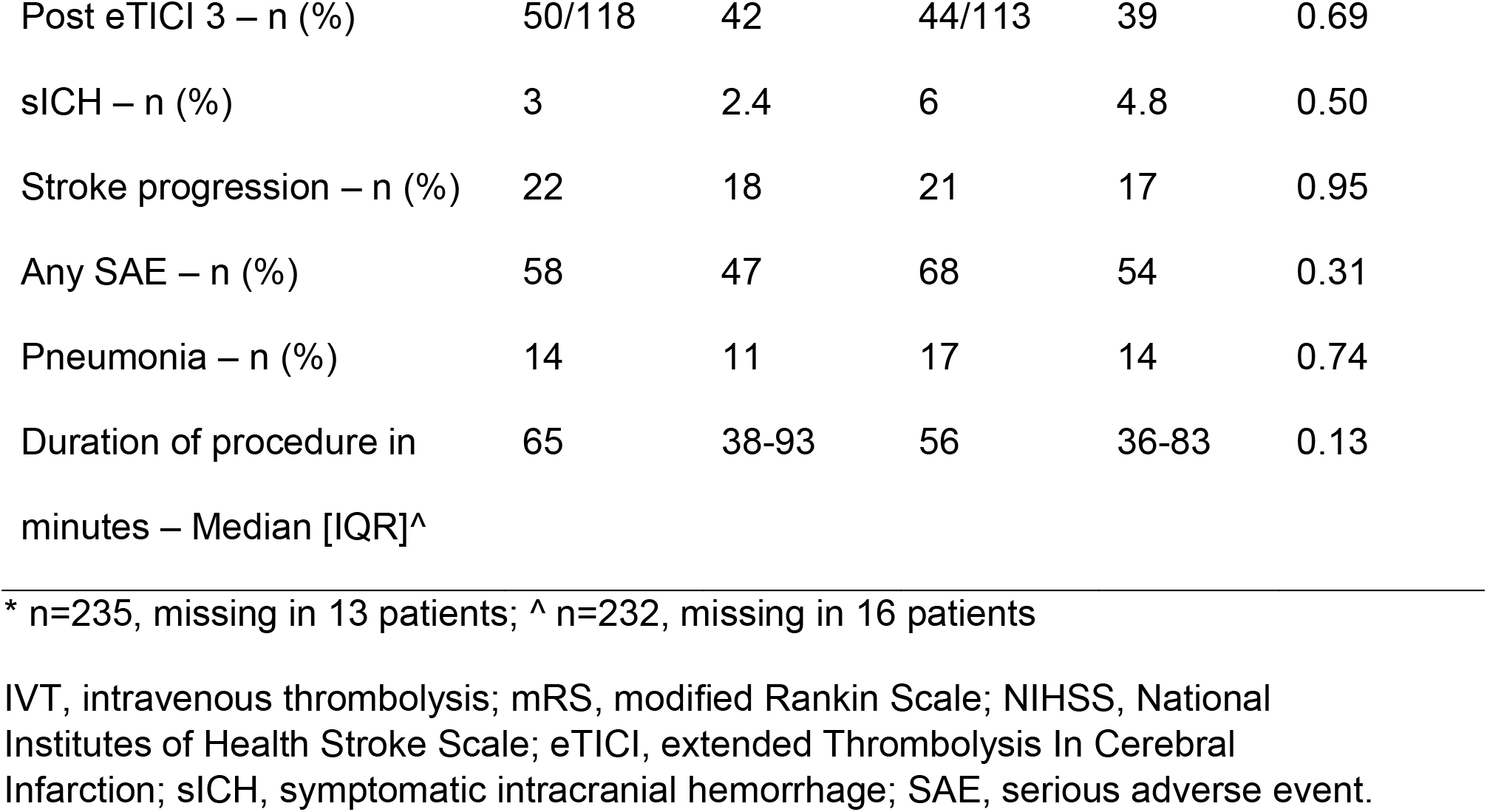

**Table 3.**

| Patients treated without IVT<br>as first modality | EE | Unadjusted<br>(95% CI) | Adjusted<br>(95% CI) |
| --- | --- | --- | --- |
| mRS at 90 days* | cOR | 0.99 (0.62 – 1.57) | 1.04 (0.61 – 1.76) |
| mRS 0-2 at 90 days | OR | 1.09 (0.64 – 1.84) | 1.02 (0.54 – 1.93) |
| mRS 0-3 at 90 days | OR | 0.84 (0.51 – 1.39) | 0.80 (0.43 – 1.49) |
| Mortality at 90 days | OR | 0.93 (0.56 – 1.55) | 0.93 (0.50 – 1.74) |
| Post EVT eTICI | cOR | 0.76 (0.48 – 1.19) | 0.78 (0.47 – 1.28) |
| Successful recanalization<br>(eTICI ≥ 2B) | OR | 0.67 (0.38 – 1.19) | 0.70 (0.37 – 1.32) |
| Excellent recanalization<br>(eTICI 3) | OR | 0.79 (0.47 – 1.33) | 0.74 (0.41 – 1.34) |
| Any serious adverse event | OR | 1.34 (0.81 – 2.21) | 1.44 (0.82 – 2.54) |
| Symptomatic ICH | OR | 2.02 (0.49 – 8.31) | 1.65 (0.33 – 8.35) |
| Stroke progression | OR | 0.93 (0.48 – 1.80) | 0.98 (0.45 – 2.10) |
| Pneumonia | OR | 1.23 (0.57 – 2.62) | 1.09 (0.47 – 2.52) |
| First-attempt successful<br>recanalization | OR | 0.98 (0.50 – 1.94) | 1.26 (0.57 – 2.77) |
| NIHSS at 24-48 hours | % | -6.8 (-30 – 25) | -1.5 (-26 – 31) |
| Procedure time | % | -13 (-26 – 2.2) | -13 (-27 – 3.7) |

\*: shift towards a better functional outcome on the full scale.
mRS, modified Rankin Scale; eTICI, extended Thrombolysis In Cerebral Infarction; ICH, intracranial hemorrhage; NIHSS, National Institutes of Health Stroke Scale.

### Clinical outcome

There was no significant difference in the mRS score at 90 days between patients treated with IVT and without IVT (acOR:1.04, 95%CI:0.61-1.76). Also, no differences were seen in mortality and favorable functional outcome at 90 days, aOR:0.93 (95%CI:0.50-1.74), and aOR:0.80 (95%CI:0.43-1.49) respectively.

### Technical outcome

Although patients treated with IVT prior to EVT had an on average shorter procedure time (56 vs 65 minutes, p =0.13), no significant differences were seen in the adjusted regression analysis after transforming the data (-13%, 95%CI:-27 – 3.7). Additionally, no differences were seen in first-attempt successful recanalization rates and successful recanalization rates (aOR:1.26, 95%CI:0.57-2.77 and aOR:0.70, 95%CI:0.37-1.32 respectively).

### Safety outcome

In 47% of the patients treated without IVT and in 54% with IVT prior to EVT any SAE occurred (p=0.31). Symptomatic ICH was twice as often seen in patients treated with IVT prior to EVT (4.8% versus 2.4%), however, this difference was not statistically significant in regression analysis (aOR:1.65, 95%CI:0.33 – 8.35).

### Subgroup analysis

There was a significant interaction between occlusion location and IVT on the mRS score at 90 days (p<0.00). In the subgroup analyses, IVT had a negative association with mRS score at 90 days (meaning higher mRS scores) in patients with an isolated posterior cerebral artery occlusion (acOR: 0.08, 95%CI: 0.00-0.72) (Supplemental Figure 1). There was a trend towards a better functional outcome in patients with an isolated basilar artery occlusions treated with prior IVT (acOR: 2.28, 95%CI: 0.95-5.49).

## Discussion

In this study, the use of IVT prior to EVT in patients with a posterior circulation occlusion did not lead to significant differences in clinical, technical, and safety outcomes.

Literature is scarce about the impact of IVT prior to EVT in patients with ischemic stroke due to posterior circulation occlusion. In the anterior circulation, multiple studies, including trials and registries, showed no superiority or non-inferiority in patients treated with IVT prior to EVT on functional outcome at 90 days.(2, 6, 17) No trials are performed yet on the effect of IVT prior to EVT in the posterior circulation.

In the BAOCHE, ATTENTION, and BEST trials 15%, 34%, and 27% of the patients received IVT respectively,(9, 11, 12) while 79% of patients in the BASICS trial received IVT.(10) Main reason for the difference is the treatment window. BAOCHE included patients between 6-24 hours after symptom onset, the ATTENTION up to 12 hours of estimated time of BAO (eBAO), and the BEST up to 8 hours after eBAO, while BASICS patients were included within 6 hours of eBAO. Another reason may be that BAOCHE, ATTENTION, and BEST included patients from China, where, to receive IVT, payment in advance is required.(9, 11, 12) In the Netherlands IVT is reimbursed, which may explain the higher rates of IVT in the MR CLEAN Registry.

The four above mentioned trials showed around 45% favorable functional outcome (mRS 0-3) in the EVT group. Similar results are presented in the current study, supplemental 2 gives an overview of favorable functional outcome in patients with only a BAO. Favorable functional outcome was seen in 44% in patients with BAO treated with IVT and 42% in patients not treated with IVT. Despite the lower pre-mRS in patients treated with IVT, no differences in favorable functional outcome were seen.

Subgroup analysis on occlusion location (supplemental 3) suggests that the potential benefit of IVT diminishes as the occlusion is more distally located when combined with EVT. Analyses were performed on a limited number of patients precluding strong conclusions, indicating the need for pooling data.

In 4.8% of the patients treated with IVT prior to EVT a sICH was seen. Comparable sICH rates were seen in the EVT groups of the BASICS (4.5%), ATTENTION (6%), BEST (8%), and BAOCHE (5%) trials.(9–12) However, these EVT groups include patients treated with and without IVT prior to EVT, while different sICH criteria were used.

Patients treated with IVT prior to EVT showed higher rates of early recanalization (17%) compared to patients treated without IVT prior to EVT (5.7%). These higher rates did not lead to differences in clinical outcome. The clinical outcome measure (mRS score at 90 days) may not be optimal to detect small differences in clinical outcomes, and a more sensitive outcome measure may be needed.

Our study has limitations. First of all, patients who recanalized after treatment with IVT alone were not included in the MR CLEAN Registry, since only EVT treated patients are included. This selection bias causes an underestimation of the effect of IVT prior to EVT in patients with a BAO. Secondly, during the MR CLEAN Registry, many patients with basilar artery occlusion were included (when eligible) in the BASICS trial, causing also potential selection bias. However, this selection bias was probably limited, since a previous publication showed similar favorable functional outcome in patients treated within the MR CLEAN Registry compared to the BASICS trial.(18) Thirdly, our registry based on real-world data has the limitations of a non-randomized study: use of IVT was left to the treating physician and the numbers are small. In the American Heart Association (AHA) guidelines IVT is contraindicated in some patients using anticoagulation and with high systolic blood pressures(19); to minimize this effect analyses were adjusted for these potential confounders. Finally, in this study, thrombus characteristics, such as the length of the occlusion and thrombus density, were not taken into account in the analysis. However, the impact of these characteristics seems to be limited.(20)

## Conclusion

We found no significant differences in clinical, technical and safety outcomes between patients with a large vessel occlusion in the posterior circulation treated with or without IVT prior to endovascular therapy. Our results are in line with the literature about the anterior circulation.

## Data Availability

Source data will not be made available because of legislative issues on patient privacy. Detailed statistical analyses and analytic methods will be made available on reasonable request to the corresponding author.

## Acknowledgments

We thank all the investigators of the MR CLEAN (Multicenter Randomized Controlled Trial of Endovascular Treatment for Acute Ischemic Stroke in the Netherlands) Registry for their effort and contributions.

## MR CLEAN Registry investigators

### Executive committee

Diederik W.J. Dippel^1^; Aad van der Lugt^2^; Charles B.L.M. Majoie^3^; Yvo B.W.E.M. Roos^4^; Robert J. van Oostenbrugge^5^; Wim H. van Zwam^6^; Jelis Boiten^14^; Jan Albert Vos^8^

### Study coordinators

Ivo G.H. Jansen^3^; Maxim J.H.L. Mulder^1,2^ ;Robert-Jan B. Goldhoorn^5,6^; Kars C.J. Compagne^2^; Manon Kappelhof^3^; Josje Brouwer^4^; Sanne J. den Hartog^1,2,40^; Wouter H. Hinsenveld ^5,6^

### Local principal investigators

Diederik W.J. Dippel^1^; Bob Roozenbeek^1^; Aad van der Lugt^2^; Charles B.L.M. Majoie^3^; Yvo B.W.E.M. Roos^4^; Bart J. Emmer^3^; Jonathan M. Coutinho^4^; Wouter J. Schonewille^7^; Jan Albert Vos^8^; Marieke J.H. Wermer^9^; Marianne A.A. van Walderveen^10^; Adriaan C.G.M. van Es^10^; Julie Staals^5^; Robert J. van Oostenbrugge^5^; Wim H. van Zwam^6^; Jeannette Hofmeijer^11^; Jasper M. Martens^12^; Geert J. Lycklama à Nijeholt^13^; Jelis Boiten^14^; Sebastiaan F. de Bruijn^15^; Lukas C. van Dijk^16^; H. Bart van der Worp^17^; Rob H. Lo^18^; Ewoud J. van Dijk^19^; Hieronymus D. Boogaarts^20^; J. de Vries^22^; Paul L.M. de Kort^21^; Julia van Tuijl^21^ ; Jo P. Peluso^26^; Puck Fransen^22^; Jan S.P. van den Berg^22^; Boudewijn A.A.M. van Hasselt^23^; Leo A.M. Aerden^24^; René J. Dallinga^25^; Maarten Uyttenboogaart^28^; Omid Eschgi^29^; Reinoud P.H. Bokkers^29^; Tobien H.C.M.L. Schreuder^30^; Roel J.J. Heijboer^31^; Koos Keizer^32^; Lonneke S.F. Yo^33^; Heleen M. den Hertog^22^; Emiel J.C. Sturm^35^; Paul J.A.M. Brouwers^34^

### Imaging assessment committee

Charles B.L.M. Majoie^3^(chair); Wim H. van Zwam^6^; Aad van der Lugt^2^; Geert J. Lycklama à Nijeholt^13^; Marianne A.A. van Walderveen^10^; Marieke E.S. Sprengers^3^; Sjoerd F.M. Jenniskens^27^; René van den Berg^3^; Albert J. Yoo^38^; Ludo F.M. Beenen^3^; Alida A. Postma^6^; Stefan D. Roosendaal^3^; Bas F.W. van der Kallen^13^; Ido R. van den Wijngaard^13^; Adriaan C.G.M. van Es^10^; Bart J. Emmer^,3^; Jasper M. Martens^12^; Lonneke S.F. Yo^33^; Jan Albert Vos^8^; Joost Bot^36^; Pieter-Jan van Doormaal^2^; Anton Meijer^27^; Elyas Ghariq^13^; Reinoud P.H. Bokkers^29^; Marc P. van Proosdij^37^; G. Menno Krietemeijer^33^; Jo P. Peluso^26^; Hieronymus D. Boogaarts^20^; Rob Lo^18^; Wouter Dinkelaar^41^; Auke P.A. Appelman^29^; Bas Hammer^16^; Sjoert Pegge^27^; Anouk van der Hoorn^29^; Saman Vinke^20^; Sandra Cornelissen^2^; Christiaan van der Leij^6^; Rutger Brans^6^

### Writing committee

Diederik W.J. Dippel^1^(chair); Aad van der Lugt^2^; Charles B.L.M. Majoie^3^; Yvo B.W.E.M. Roos^4^; Robert J. van Oostenbrugge^5^; Wim H. van Zwam^6^; Geert J. Lycklama à Nijeholt^13^; Jelis Boiten^14^; Jan Albert Vos^8^; Wouter J. Schonewille^7^; Jeannette Hofmeijer^11^; Jasper M. Martens^12^; H. Bart van der Worp^17^; Rob H. Lo^18^

### Adverse event committee

Robert J. van Oostenbrugge^5^(chair); Jeannette Hofmeijer^11^; H. Zwenneke Flach^23^

### Trial methodologist

Hester F. Lingsma^40^

### Research nurses / local trial coordinators

Naziha el Ghannouti^1^; Martin Sterrenberg^1^; Wilma Pellikaan^7^; Rita Sprengers^4^; Marjan Elfrink^11^; Michelle Simons^11^; Marjolein Vossers^12^; Joke de Meris^14^; Tamara Vermeulen^14^; Annet Geerlings^19^; Gina van Vemde^22^; Tiny Simons^30^; Gert Messchendorp^28^; Nynke Nicolaij^28^; Hester Bongenaar^32^; Karin Bodde^24^; Sandra Kleijn^34^; Jasmijn Lodico^34^; Hanneke Droste^34^; Maureen Wollaert^5^; Sabrina Verheesen^5^; D. Jeurrissen^5^; Erna Bos^9^; Yvonne Drabbe^15^; Michelle Sandiman^15^; Nicoline Aaldering^11^; Berber Zweedijk^17^; Jocova Vervoort^21^; Eva Ponjee^22^; Sharon Romviel^19^; Karin Kanselaar^19^; Denn Barning^10^

### Clinical/imaging data aquisition

Esmee Venema^40^; Vicky Chalos^1,40^; Ralph R. Geuskens^3^; Tim van Straaten^19^; Saliha Ergezen^1^; Roger R.M. Harmsma^1^; Daan Muijres^1^; Anouk de Jong^1^; Olvert A. Berkhemer^1,3,6^; Anna M.M. Boers^3,39^; J. Huguet^3^; P.F.C. Groot^3^; Marieke A. Mens^3^; Katinka R. van Kranendonk^3^; Kilian M. Treurniet^3^; Manon L. Tolhuisen^3,39^; Heitor Alves^3^; Annick J. Weterings^3^; Eleonora L.F. Kirkels^3^; Eva J.H.F. Voogd^11^; Lieve M. Schupp^3^; Sabine L. Collette^28,29^; Adrien E.D. Groot^4^; Natalie E. LeCouffe^4^; Praneeta R. Konduri^39^; Haryadi Prasetya^39^; Nerea Arrarte-Terreros^39^; Lucas A. Ramos^39^ ; Nikki Boodt^1,2,40^; Anne F.A.V Pirson^5^; Agnetha A.E. Bruggeman^3^

### List of affiliations

Department of Neurology^1^, Radiology^2^, Public Health^40^, Erasmus MC University Medical Center;

Department of Radiology and Nuclear Medicine^3^, Neurology^4^, Biomedical Engineering & Physics^39^, Amsterdam UMC, University of Amsterdam, Amsterdam; Department of Neurology^5^, Radiology^6^, Maastricht University Medical Center and Cardiovascular Research Institute Maastricht (CARIM);

Department of Neurology^7^, Radiology^8^, Sint Antonius Hospital, Nieuwegein,;

Department of Neurology^9^, Radiology^10^, Leiden University Medical Center;

Department of Neurology^11^, Radiology^12^, Rijnstate Hospital, Arnhem;

Department of Radiology^13^, Neurology^14^, Haaglanden MC, the Hague;

Department of Neurology^15^, Radiology^16^, HAGA Hospital, the Hague;

Department of Neurology^17^, Radiology^18^, University Medical Center Utrecht;

Department of Neurology^19^, Neurosurgery^20^, Radiology^27^, Radboud University Medical Center, Nijmegen;

Department of Neurology^21^, Radiology^26^, Elisabeth-TweeSteden ziekenhuis, Tilburg;

Department of Neurology^22^, Radiology^23^, Isala Klinieken, Zwolle;

Department of Neurology^24^, Radiology^25^, Reinier de Graaf Gasthuis, Delft;

Department of Neurology^28^, Radiology^29^, University Medical Center Groningen;

Department of Neurology^30^, Radiology^31^, Atrium Medical Center, Heerlen;

Department of Neurology^32^, Radiology^33^, Catharina Hospital, Eindhoven;

Department of Neurology^34^, Radiology^35^, Medisch Spectrum Twente, Enschede;

Department of Radiology^36^, Amsterdam UMC, Vrije Universiteit van Amsterdam, Amsterdam;

Department of Radiology^37^, Noordwest Ziekenhuisgroep, Alkmaar;

Department of Radiology^38^, Texas Stroke Institute, Texas, United States of America;

Department of Radiology^41^, Albert Schweitzer Hospital, Dordrecht.

## Sources of Funding

The MR CLEAN Registry (Multicenter Randomized Clinical Trial of Endovascular Treatment of Acute Ischemic Stroke) was partly funded by *Stichting Toegepast Wetenschappelijk instituut voor Neuromodulatie* (TWIN), Erasmus MC University Medical Center, Maastricht University Medical Center, and Amsterdam University Medical Center.

## Disclosures

CBLMM reports grants from the Netherlands Cardiovascular Research Initiative, an initiative of the Dutch Heart Foundation, European Commission, Healthcare Evaluation Netherlands, and Stryker (all paid to institution); and is a (minority interest) shareholder of Nicolab. BJE reports grants from leading the Change Healthcare Evaluation program, and TKI-PPP Grant Topsector lifesciences (all paid to institution); and participates as a representative of the UEMS Neuroradiology Dutch, and as Board member of the Dutch Society of Radiology. P-JvD reports consulting fees from Stryker, Siemens, and Stryker (all paid to institution); participates in the advisory board of DX Medical solutions; and is shareholder of DX Medical Solutions. AJY reports grants from Medtronic, Cerenovus, Penumbra, Stryker, and Genentech; holds stock options in Nico-lab; and is a consultant for Vesalio, Cerenovus, Penumbra, and Philips, all outside the submitted work. WHvZ reports speaker fees from Stryker, Cerenovus, and Nicolab, and consulting fees from Philips (all paid to institution); participated in the advisory boards of WeTrust (Philips) and ANAIS (Anaconda) (all paid to institution); and participated in the advisory boards of InEcxtremis (CHU Montpellier, Montpellier, France) and DISTAL (University Hospital Basel, Basel, Switzerland), studies for which no payments were received. All other authors declare no competing interests.

## Notes

### Author Declarations

The MR CLEAN Registry study protocol was evaluated by the medical ethics committee of the Erasmus University Medical Center and permission was granted to carry out the study as a registry. The need for obtaining informed consent was waived (MEC-2014-235).

